# Hormone Use among Young People with Gender Incongruence in Norway: A Nationwide Register Study

**DOI:** 10.64898/2026.04.07.26349505

**Authors:** Ove Øyås, Per Magnus, Cecilie Bjertness Nyquist, Are Hugo Pripp, Sandra Dis Steintorsdottir, Anne Wæhre

**Affiliations:** Oslo Centre for Biostatistics and Epidemiology, Oslo University Hospital, Oslo, Norway; Centre for Fertility and Health, Norwegian Institute of Public Health, Oslo, Norway; Division of Paediatric and Adolescent Medicine, Department of Child and Adolescent Mental Health in Hospitals, Oslo University Hospital, Oslo, Norway; Department of Gender Identity Assessment, Norwegian National Center for Gender Incongruence, Oslo University Hospital, Oslo, Norway; Department of Endocrinology, Morbid Obesity and Preventive Medicine, Oslo University Hospital, Oslo, Norway; Department of Clinical Science, University of Bergen, Bergen, Norway; Department of Internal Medicine, Haukeland University Hospital, Bergen, Norway; Division of Paediatric and Adolescent Medicine, Institute of Clinical Medicine, Faculty of Medicine, University of Oslo, Oslo, Norway

**Keywords:** Gender incongruence, gender-affirming hormone therapy, puberty blockers, young people

## Abstract

**Introduction:** The aim of this study was to determine the annual age- and sex-specific prevalence of gender-affirming hormone and puberty blocker use among young people with a gender incongruence (GI) diagnosis in Norway.

**Methods:** We integrated data from multiple Norwegian national registers to perform a nationwide register-based study of individuals with known sex assigned at birth who were born in the period 1975–2017 and resident in Norway for all or part of the period 2008–2022. We first calculated the annual age- and sex-specific incidence of GI diagnoses in the population. Then, we calculated the annual age- and sex-specific prevalence of androgen, estrogen, and puberty blocker use among individuals with a GI diagnosis who were under age 25 (for androgens and estrogens) or 18 (for puberty blockers) in the year that they collected the prescription.

**Results:** The incidence of GI diagnoses has increased among youth in Norway, most notably since 2015 and with the largest increase among adolescents assigned female at birth. The prevalence of feminizing and masculinizing hormone therapy has increased in this period as well, mainly among adolescents and young adults. The prevalence of puberty blocker use is mostly low but has increased since 2015, especially in recent years among adolescents assigned male at birth.

**Conclusion:** The prevalence of gender-affirming hormone and puberty blocker use has increased among young people with a GI diagnosis in Norway, mainly among adolescents and young adults, concurrently with an increase in the incidence of GI diagnoses.

## Introduction

Gender incongruence (GI) refers to a persistent mismatch between an individual’s experienced gender and their sex assigned at birth, often associated with distress or impairment described as gender dysphoria (GD) (1, 2). Medical interventions such as puberty suppression and gender-affirming hormone therapy (GAHT) can be offered to youth with GI to align physical development with gender identity (3, 4). GAHT consists of estrogen replacement and an anti-androgen for individuals assigned male at birth (AMAB) and androgen replacement for individuals assigned female at birth (AFAB). Puberty blockers such as gonadotropin-releasing hormone (GnRH) analogs suppress the production of sex hormones and are prescribed to halt the progression of puberty once it has begun.

Internationally, referrals of youth to gender identity services have increased over the past decades, particularly among AFAB individuals compared to AMAB individuals (5, 6). This has spurred interest in understanding the use of puberty blockers and gender-affirming hormones in young people with GI (7). Clinical guidelines differ in their recommendations, with international standards of care emphasizing careful, individualized assessment for gender-affirming treatment (3) while some national authorities have restricted hormonal treatment of minors pending stronger evidence (8). Little is known, however, about who receives such treatment, at what age, and how these patterns have evolved over time among youth diagnosed with GI. Such knowledge is essential for equitable care and effective health policy.

Recent nationwide register studies from across Scandinavia have begun to explore these trends. In Denmark, Glintborg et al. (9) examined prescription patterns of GAHT from 2000 to 2018. A Swedish study by Indremo et al. (10) documented a rise in GD diagnoses between 2004 and 2015, especially among individuals aged 10–30. However, these studies do not capture the most recent decade or offer detailed age- and sex-specific data on diagnosis and hormone treatment prevalence.

In Norway, the Norwegian National Center for Gender Incongruence (NCGI) at Oslo University Hospital (OUH) conducts the assessment and medical and surgical treatment of GI in youth. Gender-affirming medical and surgical treatment for adults has been provided at OUH since 1979. The exact year when the service began assessing children and adolescents with GI is unclear, but referrals of minors were documented during the early 2000s. During this period, puberty suppression was gradually introduced into clinical practice at the NCGI, drawing on the Dutch treatment approach (11), with subsequent clinical practice also informed by international guidelines (3, 4).

Today, children and adolescents are referred to the centralized child and adolescent unit at OUH, while young adults are seen at the affiliated adult clinic. Diagnoses of GI may be made in various parts of the specialist health services, including local psychiatric outpatient settings, but according to Norwegian guidelines, all medical and surgical treatments should be initiated at the national service (12). In practice, some medical treatment may have been initiated by decentralized providers outside the national service, but the number of such providers is not centrally recorded. Puberty blockers can be offered from early puberty (Tanner stages 2–3) following assessment, and GAHT is generally initiated from age 16, although in previous years some individuals might have started treatment at an earlier age.

The centralized health care model makes Norway well-suited for register-based analyses of treatment patterns. Each resident is assigned a unique personal identification number, which allows linkage across national registers. Because all publicly funded health care contacts and prescriptions are recorded, we can capture comprehensive information on diagnosis and treatment at the national level. While earlier register studies from Denmark and other Nordic countries have examined either GI diagnoses or GAHT, no nationwide studies to date have investigated age- and sex-specific trends in both incidence of GI diagnoses and prevalence of puberty blocker use and GAHT among youth.

In this study, we integrated data from several Norwegian national registers to estimate (1) the annual incidence of GI diagnoses among individuals under age 25 and (2) the annual prevalence of GAHT and puberty blocker use among individuals diagnosed with GI (under age 25 for GAHT and under age 18 for puberty blockers), stratified by age and sex assigned at birth. Our results provide a national baseline for understanding treatment uptake and timing.

## Methods

### Study population

We performed a nationwide cross-sectional register-based study using data from Norwegian national registers. Data were obtained for all individuals born in the period 1975–2017 who were resident in Norway for all or part of the period 2008–2022. Individuals born after 2017 were not included because GI diagnoses are rarely given to children under age four, and individuals who resided in Norway only before the Norwegian Patient Registry was established in 2008 could not contribute data. To enable sex-specific analyses, the study population was defined as individuals with known sex assigned at birth, as reported in the Medical Birth Registry of Norway. All analyses were restricted to individuals under age 25, or under age 18 for puberty blockers, in the year of diagnosis or prescription collection.

### Data sources

We obtained year of birth from Statistics Norway for all 3,226,906 individuals meeting the birth and residency criteria. Of these, 2,405,222 individuals (75%) had a registered sex assigned at birth (male/female) in the Medical Birth Registry of Norway and were included in the study population. The 821,684 individuals (25%) without registered sex assigned at birth were largely people born outside Norway, who are not covered by this registry. From the Norwegian Patient Registry, we obtained GI diagnosis codes and year of diagnosis recorded by the secondary care services between 2008 and 2022, specifically for the following codes from ICD-10 and the Norwegian modification ICD-10-NO (Z-codes introduced in 2021 to align with ICD-11 categories): F64.0 (transsexualism), F64.2 (gender identity disorder in childhood), F64.8 (other gender identity disorders), F64.9 (unspecified gender identity disorder), Z76.80 (gender incongruence in adolescents and adults), Z76.81 (gender incongruence in childhood), and Z76.89 (unspecified gender incongruence). Prescriptions of GAHT and puberty blockers collected from pharmacies between 2004 and 2022, including year of collection, were provided by the Norwegian Prescribed Drug Registry, specifically for the ATC codes G03B (androgens), G03C (estrogens), and L02AE02 (leuprorelin). All datasets included an anonymized personal identifier enabling linkage across registers. Age at diagnosis and drug prescription collection were calculated as year of diagnosis or year of drug collection minus year of birth.

### Incidence of gender incongruence diagnoses

We defined a GI diagnosis as any occurrence of one or more of the ICD-10 and ICD-10-NO codes corresponding to GI. These seven codes were pooled to represent a single category of GI for all analyses. Annual incidence of GI diagnoses was calculated for each year from 2008 to 2022 as the number of individuals receiving their first-ever GI diagnosis (i.e., any one of the seven codes) divided by the number of individuals at risk that year (i.e., those without a prior GI diagnosis). We did this separately for AMAB and AFAB while also grouping by age at diagnosis into children (0–11), young adolescents (12–14), adolescents (15–17), and young adults (18–24). Incidence rates of GI diagnoses are reported per 100,000 person-years.

### Prevalence of gender-affirming hormone and puberty blocker use

For each year from 2008 to 2022, we identified individuals under age 25 who had received a GI diagnosis in the same year or earlier. Among these individuals, we calculated the annual prevalence of GAHT or puberty blocker use as the proportion who collected at least one relevant prescription that year. We refer to this as use for brevity, although collection of a prescription does not confirm that the medication was taken. GAHT was defined as androgen use for AFAB and estrogen use for AMAB, and puberty blocker use was defined as collection of leuprorelin. Leuprorelin is the only GnRH analog routinely prescribed for puberty suppression in Norway during the study period. Among children and young adolescents, it is given as monotherapy to suppress puberty, whereas among older AMAB adolescents who have started estrogen therapy it may also be given alongside estrogen as an anti-androgen. Since most AFAB and AMAB individuals will have completed puberty by age 18, individuals aged 18 years or older were excluded from all puberty blocker analyses. Analyses were limited to androgens, estrogens, and leuprorelin to ensure valid attribution of pharmacological treatment to gender-affirming care. Purely anti-androgenic agents were not included, as they are frequently prescribed for other indications and are not specific to gender-affirming care. Prevalence was stratified by age group and sex assigned at birth (as we did for incidence), and rates are reported separately for androgens, estrogens, and puberty blockers as a percentage of individuals with a GI diagnosis.

### Statistical analyses

We calculated exact 95% confidence intervals for incidence and prevalence estimates using the Clopper–Pearson method (binom.test in R). To estimate the effects of sex assigned at birth and age group on incidence of GI diagnoses and prevalence of hormone use since 2015, we fitted a negative binomial regression model to data from the period 2015–2022 (glmmTMB in R). We chose this model over a Poisson model due to overdispersion. We used the number of individuals who received their first GI diagnosis or collected a hormone as response (offset by total group size) and sex assigned at birth, age group, and their interaction as predictors. Androgens and estrogens were combined into a single model, as they are almost exclusively used by AFAB and AMAB individuals, respectively. Age groups with fewer than five events for one sex assigned at birth were excluded (children aged 0–11 for all hormones and young adolescents aged 12–14 for gender-affirming hormones). We used the fitted models to perform pairwise comparisons of incidence and prevalence between age groups and between AMAB and AFAB, adjusting *p*-values to control the false discovery rate using the Benjamini-Hochberg procedure (emmeans in R). All analyses were performed using R version 4.2.3.

## Results

### Annual incidence of gender incongruence has increased among youth since 2015

We first calculated the annual rate of individuals under age 25 who received their first GI diagnosis in specialist health care services between 2008 and 2022 (**Fig. 1**), separately for each sex assigned at birth and grouping individuals by age into children (0–11), young adolescents (12–14), adolescents (15–17), and young adults (18–24). Incidence of GI diagnoses was stable and comparatively low across sex and age groups (below 20 / 100,000) from 2008 until 2014, after which it started increasing (**Supplemental Table 1**). In total, 2,590 individuals in the population received at least one GI diagnosis before age 25 during this period (1,561 AFAB and 1,029 AMAB).

**Fig. 1:**
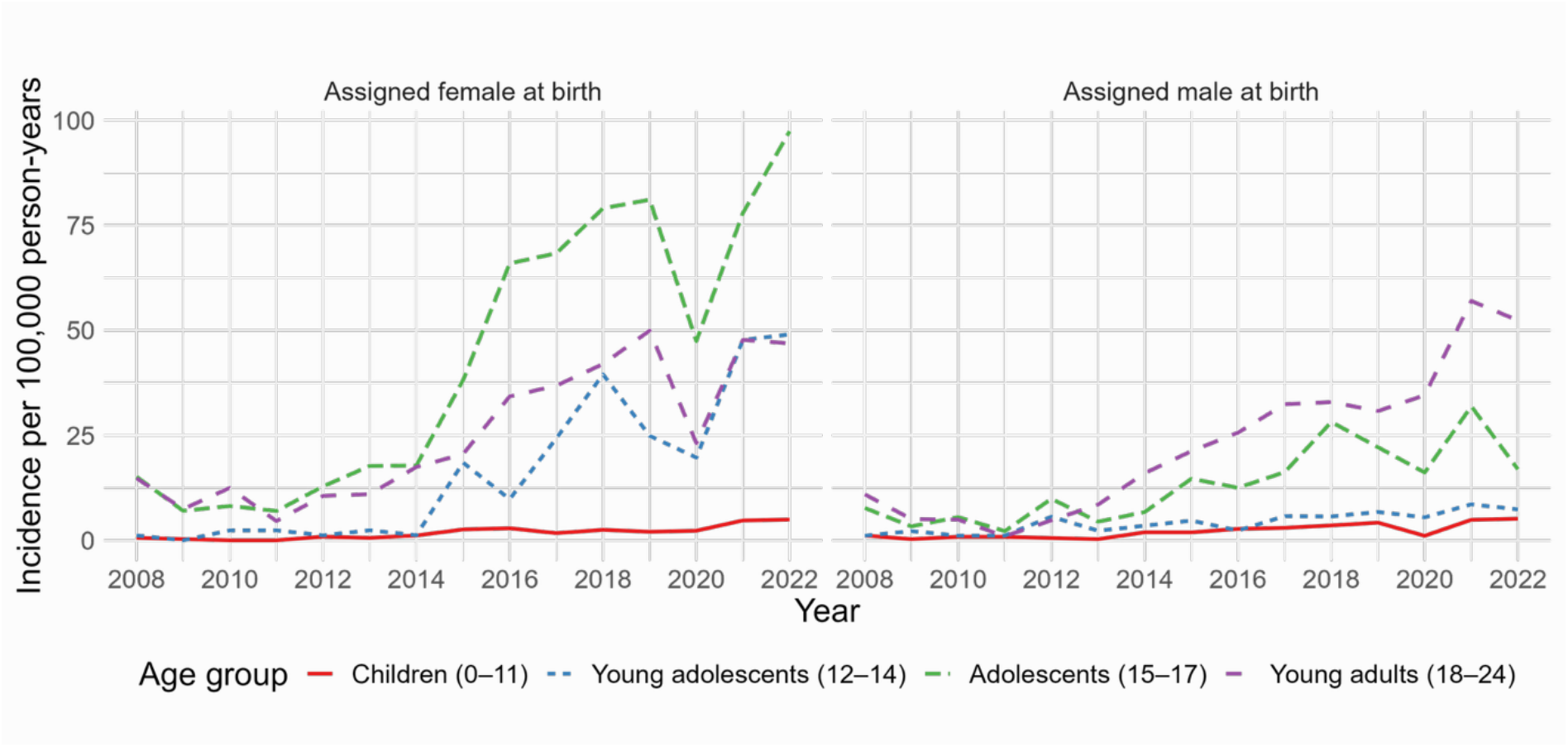
Sex-specific annual incidence of gender incongruence (GI) diagnoses in the study population, grouped by age at time of diagnosis. Seven different diagnoses were pooled to represent GI, and incidence was calculated as the number of people receiving any one of these diagnoses for the first time per 100,000 person-years within each group.

Among young adolescents and adolescents, a greater increase in incidence of GI was observed for AFAB than for AMAB. Incidence among AFAB adolescents increased sharply from 2015, rising from 18 per 100,000 in 2014 to 97 per 100,000 in 2022, approaching 1 per 1,000. Among AFAB young adolescents, the increase was more variable and reached approximately 1 per 2,000 in 2022, comparable to the incidence among AFAB young adults. The increase in incidence of GI since 2015 among young adults has been similar for AFAB and AMAB, and the same is true for the low incidence among children. There was a notable general decrease in incidence around 2020, which could be explained by the COVID-19 pandemic or a transition to new diagnosis codes that year, and this effect also appears to be greater in AFAB than in AMAB.

Considering just the absolute number of individuals who received their first GI diagnosis each year, we found similar trends as for incidence (**Supplemental Figure 1**). However, the largest increase by absolute counts has been among young adults rather than adolescents, peaking at 116 AMAB individuals in 2021 and 106 in 2022, compared to 92 and 90 AFAB individuals in the same years. Since 2015, counts have been larger for AFAB than for AMAB among both young adolescents and adolescents. Again, the increase in GI diagnoses among AFAB adolescents since 2015 is not reflected in AMAB, but more AMAB adults than AFAB adults received their first diagnosis in 2021 and 2022.

To examine these patterns more formally, we fitted a regression model to GI diagnoses given between 2015 and 2022 to estimate the effects of sex assigned at birth, age group, and their interaction on incidence in this period. Pairwise comparisons within each sex assigned at birth showed significant differences in incidence between all age groups for AMAB and between all age groups except young adolescents and young adults for AFAB (**Supplemental Table 2**). Supporting our earlier observations, incidence was significantly higher for AFAB than for AMAB among young adolescents and adolescents (**Table 1**). Because the number of diagnoses rose from very different baseline levels, comparable relative increases produced the largest absolute increases among young adults, who had the highest number of diagnoses at the start of the period.

**Table 1:**
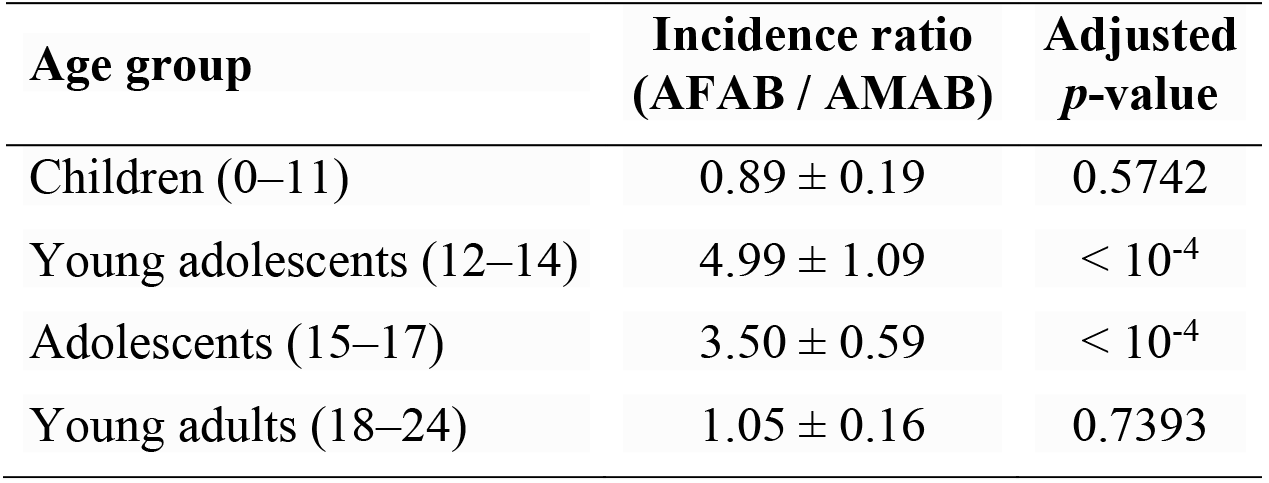
Estimated sex ratios for incidence of gender incongruence diagnoses in the study population. Ratios are shown for individuals assigned female at birth (AFAB) relative to individuals assigned male at birth (AMAB) and for each age group with standard error and *p*-value (adjusted by the Benjamini-Hochberg procedure). Estimates come from a regression model fitted to data from the period 2015–2022.

### Annual prevalence of hormone use has increased among young people with GI since 2015

Similar to incidence of GI diagnoses, we found that prevalence of hormone use has increased markedly since 2015 (**Fig. 2; Supplemental Table 3**). Comparing prevalence of hormone use between AFAB and AMAB, we found similar patterns of gender-affirming hormone use but apparent differences in the application of puberty blockers. Since 2015, the prevalence of GAHT has mainly increased among adolescents and young adults. Among AFAB young adults, prevalence of androgen use has exceeded 50% since 2021, and among AMAB young adults, prevalence of estrogen use rose to 39% in 2022. Among adolescents, the prevalence of gender-affirming hormone use in 2022 was 19% for AFAB and 15% for AMAB. Use of GAHT remained negligible among young adolescents and children.

**Fig. 2:**
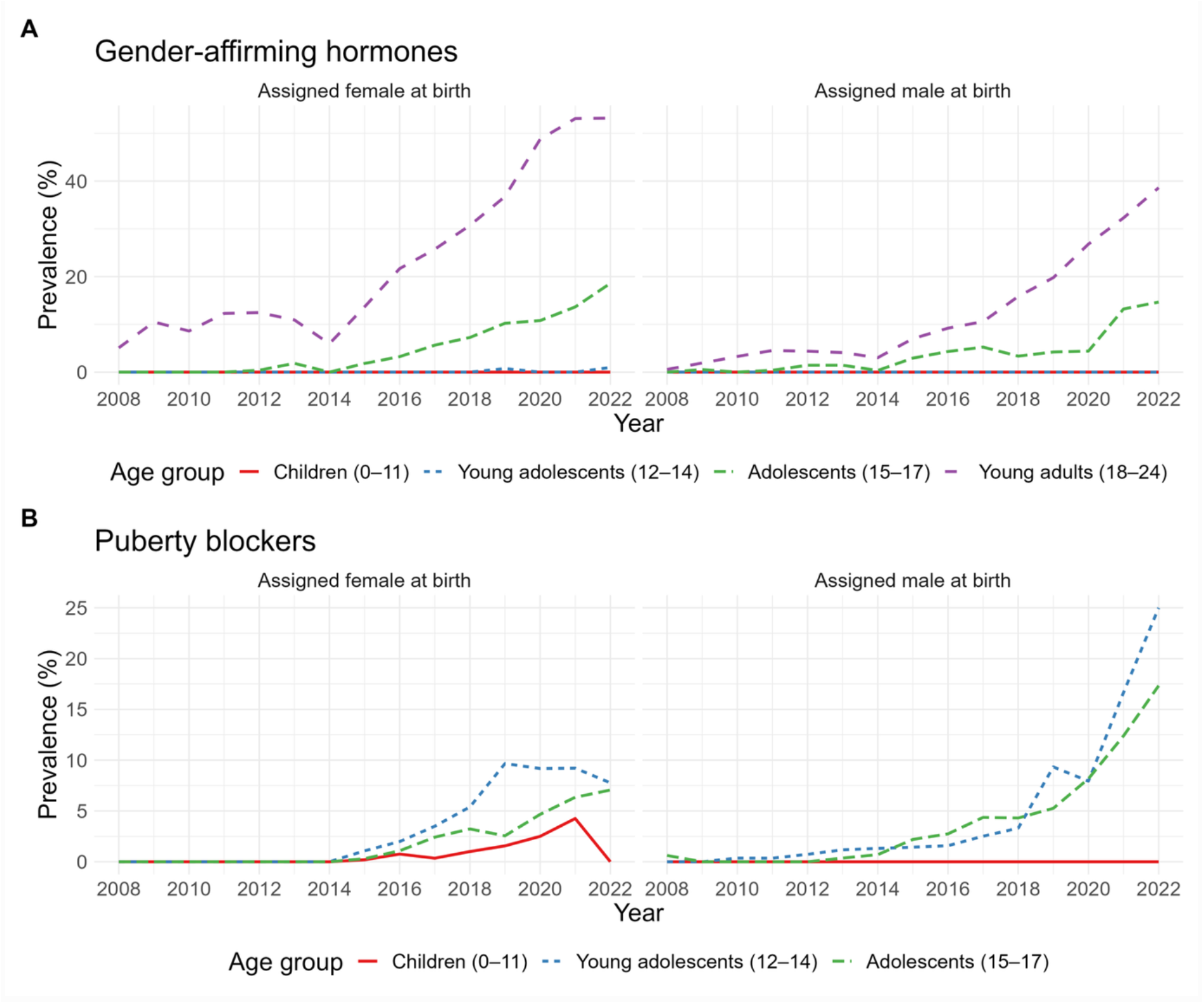
Sex-specific annual prevalence of (A) gender-affirming hormone use and (B) puberty blocker use among individuals who had received a gender incongruence diagnosis and were under age 25 (for gender-affirming hormones) or 18 (for puberty blockers) in the year that they collected the prescription, grouped by age at time of collection. Gender-affirming hormones are androgens and estrogens for individuals assigned female and male at birth, respectively, and puberty blockers are leuprorelin regardless of sex assigned at birth.

The prevalence of puberty blocker use has increased among both young adolescents and adolescents, most notably for AMAB in recent years. For AMAB, prevalence of puberty blocker use reached 25% among young adolescents and 17% among adolescents in 2022, compared with 8% and 7%, respectively, for AFAB. Among AFAB young adolescents, prevalence peaked at about 10% in 2019 and has declined slightly since. Hardly any children below age 12 received puberty blockers in 2022, although up to 4% of AFAB children with a GI diagnosis did so until 2021.

Counts of individuals receiving gender-affirming hormones followed the same pattern as prevalence for AFAB and AMAB, with steady increases in young adults receiving GAHT since 2015 (**Supplemental Figure 2**). In 2022, 436 AFAB young adults and 210 AMAB young adults received GAHT. Among AFAB young adolescents, the puberty blocker count peaked at 26 in 2019 and fell to 8 in 2022, while among AFAB adolescents it reached 20 in 2021 and 19 in 2022. For AMAB, counts generally increased, reaching 12 among young adolescents and 13 among adolescents in 2022. Despite these trends, puberty blocker use remains limited to a very small number of individuals regardless of sex assigned at birth.

Fitting regression models to hormone use prevalence data from the period 2015–2022, as we did for incidence of GI diagnoses, we found significant differences in prevalence between age groups within each sex assigned at birth for gender-affirming hormones but not for puberty blockers (**Supplemental Table 4**). The only significant difference between AFAB and AMAB was in GAHT use among young adults, with a higher prevalence for AFAB (**Table 2**).

**Table 2:** Estimated sex ratios for prevalence of gender-affirming hormone and puberty blocker use among individuals who had received a gender incongruence diagnosis and were under age 25 (for gender-affirming hormones) or 18 (for puberty blockers) in the year that they collected the prescription. Ratios are shown for individuals assigned female at birth (AFAB) relative to individuals assigned male at birth (AMAB) and for each hormone and age group with standard error and *p*-value (adjusted by the Benjamini-Hochberg procedure). Estimates come from regression models fitted to data from the period 2015–2022.

| **Hormone group** | **Age group** | **Prevalence ratio**<br>**(AFAB / AMAB)** | **Adjusted**<br>***p*-value** |
|---|---|---|---|
| Gender-affirming hormones | Adolescents (15–17) | 1.45 ± 0.42 | 0.1976 |
|  | Young adults (18–24) | 1.77 ± 0.46 | 0.0273 |
| Puberty blockers | Young adolescents (12–14) | 0.78 ± 0.30 | 0.5134 |
|  | Adolescents (15–17) | 0.51 ± 0.19 | 0.0713 |

## Discussion

This nationwide register study shows an increase in Norwegian youth diagnosed with GI and in the use of GAHT and puberty blockers since 2015, mirroring international trends. Incidence of GI diagnoses increased across youth in this period, with the steepest rise among AFAB adolescents. In parallel, the prevalence of GAHT and puberty blocker use among youth diagnosed with GI increased markedly. GAHT became more prevalent among adolescents and young adults, and puberty blocker use in adolescence remained limited but showed a more pronounced increase among AMAB in recent years. These differences by age and sex assigned at birth in both diagnosis and treatment are relevant for service planning.

The observed rise in the number of adolescents diagnosed with GI, which was largest among AFAB individuals, is consistent with national registry data from other Nordic countries (9, 10). The drivers are debated and cannot be determined from register data, but proposed factors include increased visibility of gender diversity, societal acceptance, evolving diagnostic practices, and broader access to care. Similar patterns have been described in systematic reviews (5, 6). A Swedish study (10) reported similar trends among youth aged 10–30, though it does not include the past decade, during which the largest increase in Norway has occurred. Our study adds timely age- and sex-stratified data that were previously lacking in Nordic cohorts.

Incidence of GI diagnoses among AMAB generally increased with age, without the distinct adolescent peak observed in AFAB. The number of AMAB individuals receiving their first GI diagnosis was lower than for AFAB among young adolescents and adolescents but comparable to AFAB among adults. This may reflect differences in gender identity development and care-seeking behavior. It is not clear whether this indicates a true difference in diagnosis rates between sexes assigned at birth or delayed timing, as AMAB individuals may present to care at a later age.

Treatment patterns also differed by sex assigned at birth. Among young adults, AFAB individuals received androgens at higher rates than AMAB peers received estrogen. The increase in puberty blocker use among AMAB adolescents in recent years, although prevalence did not differ significantly from AFAB, may be influenced by differences in pubertal timing and clinical considerations. In some cases, masculinization might be viewed as more difficult to reverse, particularly the development of a low-pitched voice, and clinicians may therefore be more inclined to initiate suppression in AMAB adolescents (3, 4). Additionally, AMAB individuals may use leuprorelin not only for puberty suppression but also as an anti-androgen instead of medications like cyproterone acetate or spironolactone after starting estrogen therapy. Leuprorelin use among older AMAB adolescents may partly reflect this. For AFAB youth, menstrual management can sometimes be used as an alternative to prolonged puberty suppression (3).

This study’s main strength is the use of nationwide health registers, enabling comprehensive, stratified analyses by age, sex assigned at birth, and treatment type. Limitations include broad inclusion of diagnostic codes, which may capture individuals who were not evaluated as candidates for medical treatment, and lack of data on dosage and route of administration. Because the analyses were based on annual cross-sectional counts, they cannot distinguish treatment initiation from continued treatment or describe individual treatment courses. Collection of a prescription does not confirm that the medication was taken, and hormones obtained outside Norwegian pharmacies are not captured. Diagnoses made before 2008 or outside specialist health care were not available, so some individuals classified as newly diagnosed early in the study period may have been diagnosed earlier. According to national Norwegian guidelines (12), all gender-affirming medical treatment should be initiated at the national service, yet some pharmacological treatment may have been started by decentralized providers with variable practices.

Anti-androgenic agents (e.g., bicalutamide, cyproterone acetate, spironolactone) were not included, as they are frequently prescribed for other common indications and are not specific to gender-affirming care in Norwegian clinical practice. For puberty suppression, only leuprorelin was included, as other GnRH analogs used internationally (e.g., triptorelin or histrelin) are not routinely prescribed for gender-affirming purposes in Norway. These restrictions improve specificity but may limit comparability with studies from other countries. Estimates of puberty blocker use among older adolescents, particularly AMAB individuals receiving estrogen, may partly reflect use of leuprorelin as an anti-androgen. Several groups included few individuals, which limits the precision of the estimates, and differences that were not statistically significant should not be taken as evidence of no difference. Finally, sex assigned at birth was obtained as a binary variable from the Medical Birth Registry of Norway, excluding individuals born outside the country and limiting generalizability as well as visibility of non-binary identities.

Puberty suppression and GAHT became more widely accepted and available for young people with GI over the study period, particularly after 2015. The increase in treatment prevalence that we observe likely reflects changes in clinical practice and access as well as changes in the number of young people diagnosed with GI. Our findings emphasize the need for individualized assessment, follow-up, and coordination between pediatric and adult health care services. Population-level data on timing and uptake are essential for service planning, yet the evidence base for these treatments in minors is debated and long-term outcomes of puberty suppression and GAHT initiated in adolescence remain insufficiently understood. Future research should link register and clinical data to evaluate health outcomes over time, as well as treatment types and dosages.

## Conclusion

This study provides the first national estimates of the incidence of GI diagnoses and the prevalence of puberty blocker and gender-affirming hormone use among young people with GI in Norway. Diagnoses and treatment increased after 2015, underscoring the need for continued long-term outcome research.

## Statement of Ethics

This study protocol was reviewed and approved by the Regional Committees for Medical and Health Research Ethics South East Norway (REC South East; reference number 167265). In accordance with the Norwegian Health Research Act (Section 35), REC approved the use of health data collected within the health and care services for research purposes without requiring individual consent. The requirement for written informed consent was waived by REC South East. The study was conducted in accordance with the principles of the Declaration of Helsinki.

## Data Availability Statement

Individual participant data will not be made available. Aggregated counts and rates are included in the supplemental tables. Further enquiries can be directed to the corresponding author.

## Author Contributions

Dr Ove Øyås conceptualized and designed the study, developed the methodology, implemented the computer code, analyzed and visualized the data, drafted the initial manuscript, and critically reviewed and revised the manuscript.

Drs Per Magnus, Cecilie Bjertness Nyquist, and Are Hugo Pripp conceptualized and designed the study, developed the methodology, and critically reviewed and revised the manuscript.

Dr Sandra Dis Steintorsdottir critically reviewed and revised the manuscript.

Dr Anne Wæhre acquired the funding, conceptualized and designed the study, administered the study, developed the methodology, drafted the initial manuscript, and critically reviewed and revised the manuscript.

All authors approved the final manuscript as submitted and agree to be accountable for all aspects of the work.

## References

1. World Health Organization. International statistical classification of diseases and related health problems, 11th revision (ICD-11). Geneva, Switzerland: World Health Organization; 2019.

2. Regier DA, Kuhl EA, Kupfer DJ. The DSM-5: classification and criteria changes. World Psychiatry. 2013;12(2):92–98.

3. Coleman E, Radix AE, Bouman WP, Brown GR, De Vries AL, Deutsch MB, Ettner R, Fraser L, Goodman M, Green J, Hancock AB. Standards of care for the health of transgender and gender diverse people, version 8. Int J Transgend Health. 2022;23(suppl 1):S1–259.

4. Hembree WC, Cohen-Kettenis PT, Gooren L, Hannema SE, Meyer WJ, Murad MH, Rosenthal SM, Safer JD, Tangpricha V, T’Sjoen GG. Endocrine treatment of gender-dysphoric/gender-incongruent persons: an endocrine society clinical practice guideline. J Clin Endocrinol Metab. 2017;102(11):3869–3903.

5. Thompson L, Sarovic D, Wilson P, Sämfjord A, Gillberg C. A PRISMA systematic review of adolescent gender dysphoria literature: 1) epidemiology. PLoS Glob Public Health. 2022;2(3):e0000245.

6. Taylor J, Hall R, Langton T, Fraser L, Hewitt CE. Characteristics of children and adolescents referred to specialist gender services: a systematic review. Arch Dis Child. 2024;109(suppl 2):S3–11.

7. D’hoore L, T’Sjoen G. Gender-affirming hormone therapy: an updated literature review with an eye on the future. J Intern Med. 2022;291(5):574–592.

8. Socialstyrelsen. Vård av barn och ungdomar med könsdysfori. Stockholm, Sweden: Socialstyrelsen; 2022. https://www.socialstyrelsen.se/contentassets/5e53acc447b04fe28484f9807455e61b/2022-12-8302.pdf

9. Glintborg D, Rubin KH, Kristensen SB, Lidegaard Ø, T’Sjoen G, Hilden M, Andersen MS. Gender affirming hormonal treatment in Danish transgender persons: a nationwide register-based study. Andrology. 2022;10(5):885–893.

10. Indremo M, White R, Frisell T, Cnattingius S, Skalkidou A, Isaksson J, Papadopoulos FC. Validity of the gender dysphoria diagnosis and incidence trends in Sweden: a nationwide register study. Sci Rep. 2021;11(1):16168.

11. Delemarre-van de Waal HA, Cohen-Kettenis PT. Clinical management of gender identity disorder in adolescents: a protocol on psychological and paediatric endocrinology aspects. Eur J Endocrinol. 2006;155(suppl 1):S131–137.

12. Helsedirektoratet. Kjønnsinkongruens: nasjonal faglig retningslinje. Oslo, Norway: Helsedirektoratet; 2020. https://www.helsedirektoratet.no/retningslinjer/kjonnsinkongruens

