## Supplemental Figures and Tables for "Hormone Use among Young People with Gender Incongruence in Norway: A Nationwide Register Study"

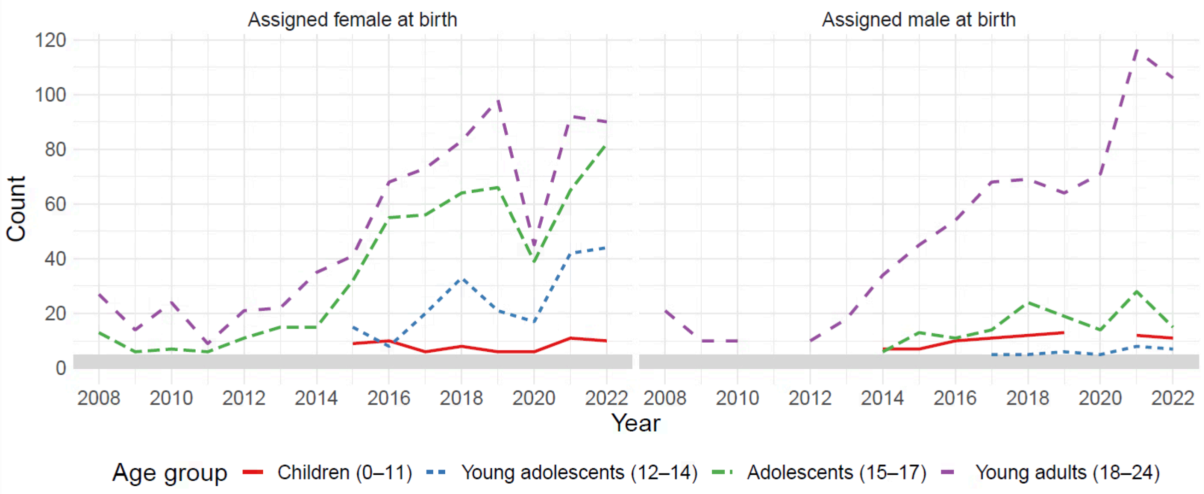


**Supplemental Figure 1:** Sex-specific annual count of individuals in the study population receiving their first gender incongruence (GI) diagnosis, grouped by age at time of diagnosis. Seven different diagnoses were pooled to represent GI, and we counted the number of people receiving any one of these diagnoses for the first time. Counts below five are not shown and fall within the shaded band.

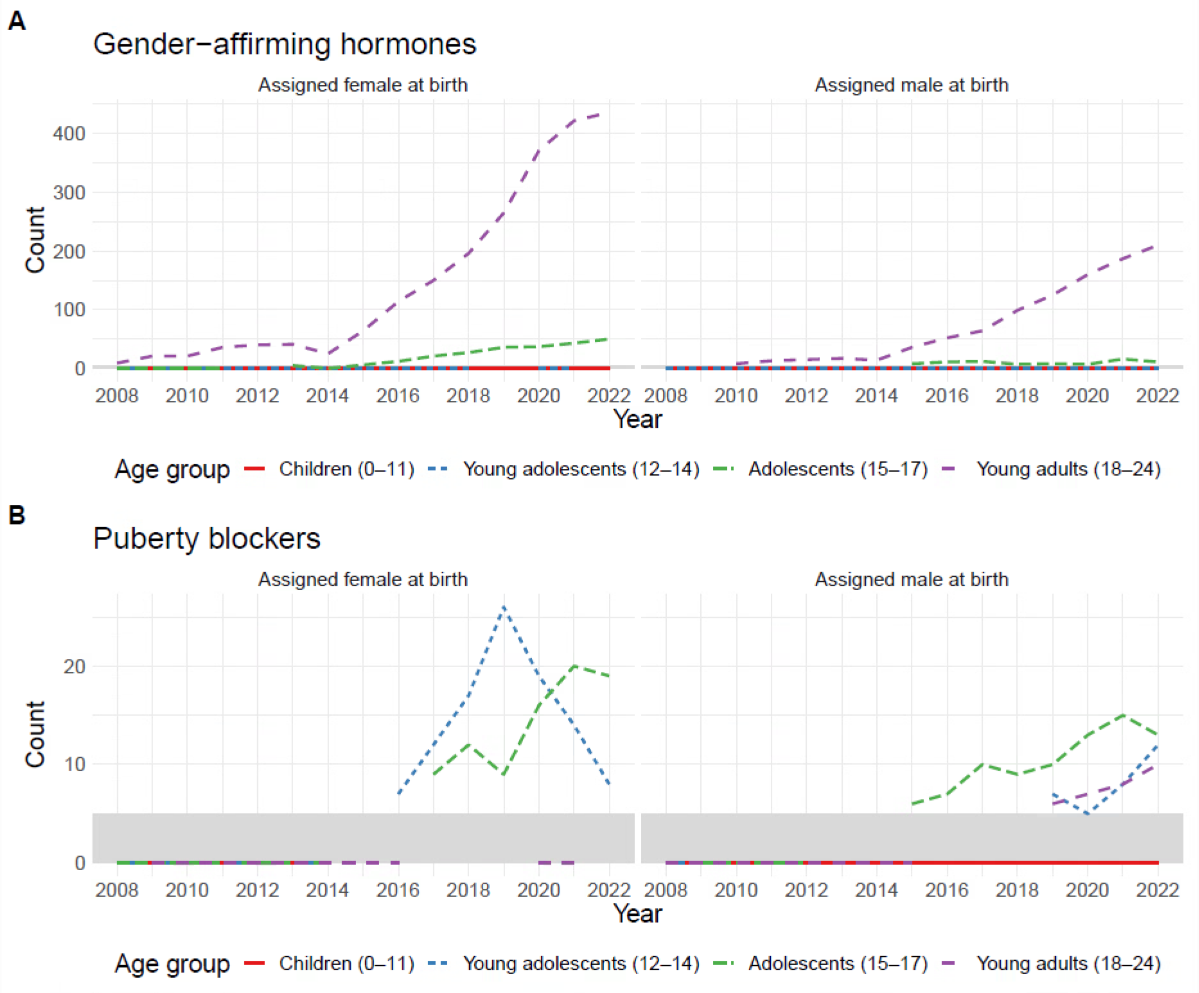


**Supplemental Figure 2:** Sex-specific annual count of individuals who collected (A) gender-affirming hormones and (B) puberty blockers who had received a gender incongruence diagnosis and were under age 25 (for gender-affirming hormones) or 18 (for puberty blockers) in the year that they collected the prescription, grouped by age at time of collection. Gender-affirming hormones are androgens and estrogens for individuals assigned female and male at birth, respectively, and puberty blockers are leuprorelin regardless of sex assigned at birth. Counts below five are not shown and fall within the shaded band.

**Supplemental Table 1:** Annual incidence of gender incongruence (GI) diagnoses in the study population in 2014 and 2022, grouped by sex assigned at birth and by age at time of diagnosis. Seven different diagnoses were pooled to represent GI. Diagnosed is the number of individuals receiving any one of these diagnoses for the first time in the given year, and incidence is that number per 100,000 person-years within each group. Counts below five are shown as “<5”.

| Sex assigned at birth | Age group | Diagnosed 2014 | Incidence 2014 | Diagnosed 2022 | Incidence 2022 |
|---|---|---|---|---|---|
| AFAB | 0–11 | <5 | 1.16 | 10 | 4.95 |
| | 12–14 | <5 | 1.22 | 44 | 49.01 |
| | 15–17 | 15 | 17.83 | 82 | 97.36 |
| | 18–24 | 35 | 17.45 | 90 | 46.87 |
| AMAB | 0–11 | 7 | 1.92 | 11 | 5.14 |
| | 12–14 | <5 | 3.48 | 7 | 7.37 |
| | 15–17 | 6 | 6.74 | 15 | 16.90 |
| | 18–24 | 34 | 15.99 | 106 | 52.34 |

**Supplemental Table 2:** Estimated age group ratios for incidence of gender incongruence diagnoses in the study population. Ratios are shown for the younger age group relative to the older age group and for each sex assigned at birth with standard error and *p*-value (adjusted by the Benjamini-Hochberg procedure). Estimates come from regression models fitted to data from the period 2015–2022.

| Sex assigned at birth | Younger group | Older group | Incidence ratio (younger / older) | Adjusted *p*-value |
|---|---|---|---|---|
| AFAB | 0–11 | 12–14 | 0.10 ± 0.02 | $< 10^{-4}$ |
| | 0–11 | 15–17 | 0.04 ± 0.01 | $< 10^{-4}$ |
| | 0–11 | 18–24 | 0.08 ± 0.01 | $< 10^{-4}$ |
| | 12–14 | 15–17 | 0.42 ± 0.07 | $< 10^{-4}$ |
| | 12–14 | 18–24 | 0.78 ± 0.12 | 0.1163 |
| | 15–17 | 18–24 | 1.84 ± 0.28 | 0.0001 |
| AMAB | 0–11 | 12–14 | 0.55 ± 0.13 | 0.0110 |
| | 0–11 | 15–17 | 0.16 ± 0.03 | $< 10^{-4}$ |
| | 0–11 | 18–24 | 0.09 ± 0.02 | $< 10^{-4}$ |
| | 12–14 | 15–17 | 0.30 ± 0.07 | $< 10^{-4}$ |
| | 12–14 | 18–24 | 0.16 ± 0.03 | $< 10^{-4}$ |
| | 15–17 | 18–24 | 0.55 ± 0.09 | 0.0004 |

**Supplemental Table 3:** Annual prevalence of gender-affirming hormone and puberty blocker use in 2014 and 2022 among individuals who had received a gender incongruence diagnosis and were under age 25 (for gender-affirming hormones) or 18 (for puberty blockers) in the year that they collected the prescription, grouped by sex assigned at birth and by age at time of collection. Gender-affirming hormones are androgens and estrogens for individuals assigned female and male at birth, respectively, and puberty blockers are leuprorelin regardless of sex assigned at birth. Collected is the number of individuals collecting at least one prescription in the given year, and prevalence is the percentage of individuals with a GI diagnosis in the same group. Counts below five are shown as "<5".

| Hormone group | Sex assigned at birth | Age group | Collected 2014 | Prevalence 2014 | Collected 2022 | Prevalence 2022 |
|---|---|---|---|---|---|---|
| Gender-affirming hormones | AFAB | 0–11 | 0 | 0.00 | 0 | 0.00 |
| | | 12–14 | 0 | 0.00 | <5 | 0.97 |
| | | 15–17 | 0 | 0.00 | 50 | 18.59 |
| | | 18–24 | 25 | 5.95 | 436 | 53.17 |
| | AMAB | 0–11 | 0 | 0.00 | 0 | 0.00 |
| | | 12–14 | 0 | 0.00 | 0 | 0.00 |
| | | 15–17 | <5 | 0.35 | 11 | 14.67 |
| | | 18–24 | 14 | 3.02 | 210 | 38.60 |
| Puberty blockers | AFAB | 0–11 | 0 | 0.00 | 0 | 0.00 |
| | | 12–14 | 0 | 0.00 | 8 | 7.77 |
| | | 15–17 | 0 | 0.00 | 19 | 7.06 |
| | AMAB | 0–11 | 0 | 0.00 | 0 | 0.00 |
| | | 12–14 | <5 | 1.31 | 12 | 25.00 |
| | | 15–17 | <5 | 0.70 | 13 | 17.33 |

**Supplemental Table 4:** Estimated age group ratios for prevalence of gender-affirming hormone and puberty blocker use among individuals who had received a gender incongruence diagnosis and were under age 25 (for gender-affirming hormones) or 18 (for puberty blockers) in the year that they collected the prescription. Ratios are shown for the younger age group relative to the older age group and for each hormone group and sex assigned at birth with standard error and *p*-value (adjusted by the Benjamini-Hochberg procedure). Estimates come from regression models fitted to data from the period 2015–2022.

| **Hormone group** | **Sex assigned at birth** | **Younger group** | **Older group** | **Prevalence ratio (younger / older)** | **Adjusted p-value** |
|---|---|---|---|---|---|
| Gender-affirming hormones | AFAB | 15–17 | 18–24 | $0.25 \pm 0.07$ | $< 10^{-4}$ |
| | AMAB | 15–17 | 18–24 | $0.30 \pm 0.09$ | $< 10^{-4}$ |
| Puberty blockers | AFAB | 12–14 | 15–17 | $1.70 \pm 0.63$ | 0.1507 |
| | AMAB | 12–14 | 15–17 | $1.12 \pm 0.44$ | 0.7779 |
